# THREE-DIMENSIONAL PRINTING OF THE MAXILLOFACIAL SEGMENT OF THE HUMAN SKULL FOR ANATOMICAL EDUCATION

**DOI:** 10.1101/2025.07.07.25329657

**Authors:** Harry James Okah, Christopher A. Ogan

## Abstract

**BACKGROUND:** Three-dimensional (3D) printing, also known as additive manufacturing or digital fabrication, is an emerging technology with diverse applications across multiple industries (Shahrubudin et. al., 2019). Despite its growing adoption, its potential in anatomical education particularly in developing countries such as Nigeria remains underexplored. This study aimed to fabricate a maxillofacial segment of the human skull using 3D printing technology to produce an anatomically accurate, ethically compliant, mobile, biosecure and cost-effective model for educational purposes.

**METHODOLOGY:** A Computer-Aided Design (CAD) model of a human skull in Stereolithography (.STL) format was processed using Ultimaker® Cura™ and Microsoft® 3D Builder™ software. The model was then printed using a Creality® Ender-3™ 3D printer with polylactic acid (PLA) filament (1.75 mm) via Fused Deposition Modeling (FDM).

**RESULTS:** The resulting model demonstrated high fidelity to anatomical structures, confirming the feasibility of 3D printing for producing biosecure, accessible, and ethically non-controversial anatomical models.

**CONCLUSION:** These findings suggest that 3D printing technology can effectively supplement traditional anatomical education, particularly in resource-limited settings where access to cadavers is limited.

## INTRODUCTION

Knowledge of maxillofacial anatomy is essential for medical and dental education, yet access to cadaveric specimens remains limited in many regions, including Nigeria. Three-dimensional (3D) printing, or additive manufacturing (AM), is a process of fabricating physical objects from digital models through successive layer deposition (Mpofu et al., 2014). Unlike subtractive manufacturing techniques (e.g., milling or drilling), 3D printing builds structures additively, allowing for complex geometries and customized designs. This technology has gained traction in various fields, including medicine, engineering, and architecture, due to its versatility and declining costs (Mpofu et al., 2014). In medical education, 3D-printed anatomical models offer a viable alternative to cadaveric specimens, which are often scarce, ethically sensitive, and associated with health risks from formaldehyde exposure (Rengier et al., 2010). Despite these advantages, the adoption of 3D printing in anatomical education particularly in developing nations like Nigeria remains limited.

While 3D printing has been extensively researched in high-income countries, its application in anatomical education within low-resource settings is underreported. The current educational landscape in many African countries often relies heavily on theory-based instruction, challenges such as limited access to cadavers, ethical concerns, and safety risks associated with traditional dissection necessitate alternative teaching tools and the integration of 3D printing introduces a practical, hands-on approach that can help students apply theoretical concepts to real-world scenarios.This study addresses this gap by demonstrating the feasibility of 3D-printed anatomical models for maxillofacial education.

The decreasing cost of 3D printers and the availability of open-source software present an opportunity to enhance anatomical training in resource-constrained environments. This study contributes to the growing body of research on 3D printing in medical education, particularly in contexts where cadaver availability is restricted.

This research highlights the potential of 3D printing to produce accurate, portable, and ethically compliant anatomical models, mitigating the limitations of cadaver-based learning.

The primary objective was to fabricate a 3D-printed maxillofacial skull segment suitable for anatomical education.

This research is limited to the application of 3D printing technology or digital fabrication in the study of Human Anatomy to produce a human skull showing maxillofacial features suitable as an anatomical model for lecture demonstration and as a learning aid.

## METHODOLOGY

### Site of Study

The study was conducted in a laboratory setting equipped with a Creality® Ender-3™ 3D printer and associated software tools.

## Materials

### Hardware

#### 3D Printer

Creality® Ender-3™ FDM printer (Shenzhen Creality 3D Technology Co., Ltd., China) with a build volume of 220 × 220 × 250 mm, positional precision of ±0.1 mm, and maximum print speed of 180 mm/s (operational speed: 30–60 mm/s). The printer features a single extruder with a maximum nozzle temperature of 250°C and a heated bed (<100°C), compatible with Windows, Mac, and Linux operating systems.

#### Filament

Polylactic acid (PLA) filament (1.75 mm diameter, natural color).

#### Computer System

HP Pavilion Gaming laptop (Intel® Core™ i5-10300H processor, NVIDIA GeForce GTX 1650, 8GB DDR4 RAM, 256GB SSD).

#### Power Supply

Sumec Firman 3000W portable generator for uninterrupted power.

#### Accessories

USB flash drive for file transfer, digital calipers for dimensional verification.

### Software

#### 3D Model

Anatomically accurate human skull model in STL format (purchased from TurboSquid®).

#### Slicing Software

Ultimaker Cura™ (v4.8, Ultimaker BV, Netherlands).

#### Segmentation Software

Microsoft 3D Builder™ (v16.0.30214.0, Microsoft Corp., USA).

### Methods

#### 1. Printer Setup and Calibration

– The Ender-3 printer was assembled according to manufacturer specifications.
– Calibration procedures included:
– Bed leveling.
– Test prints to verify dimensional accuracy.
– Nozzle temperature and extrusion rate optimization.

#### 2. Model Preparation

##### Model Acquisition

A high-resolution (0.1 mm tolerance) skull model was obtained in STL format.

**<H5>Slicing Parameters:**

– Layer height: 0.2 mm.
– Wall thickness: 1.2 mm.
– Infill density: 20% (honeycomb pattern).
– Support structures: Tree-type (density: 15%).
– Print temperature: 200°C (nozzle), 60°C (bed).

##### Model Segmentation

The skull was divided into maxillofacial and mandibular components using Boolean operations in 3D Builder™ to accommodate the printer’s build volume.

#### 3. Printing Process

– Pre-printing Checks
Filament feeding system verification.
Nozzle cleaning.
Bed adhesion treatment (glue stick application).

#### Printing Parameters

Print speed: 50 mm/s.
Cooling: 100% fan speed after the first layer.
Retraction: 6 mm at 25 mm/s.

#### Print Execution

Maxillofacial segment: 11 hours 48 minutes.

### 4. Post-Processing

– Support structure removal.
– Surface finishing with 400-grit sandpaper.
– Component alignment verification.
– Bonding with cyanoacrylate adhesive.

## RESULTS

The printed maxillofacial segment (Fig. 1) measured **26.67 × 32.39 mm** and accurately reproduced key anatomical features, including:

– Orbital margins.
– Zygomatic processes.
– Nasal conchae.
– Palatine sutures.
– Alveolar processes.

**Fig. 1.**
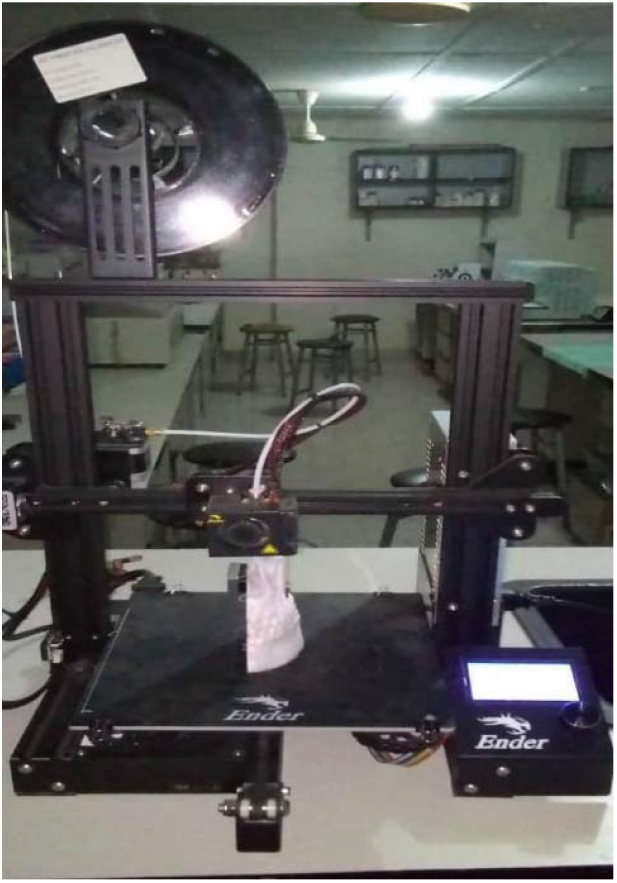
Ender-3 3D printer (printing a model).

**Fig. 2.**
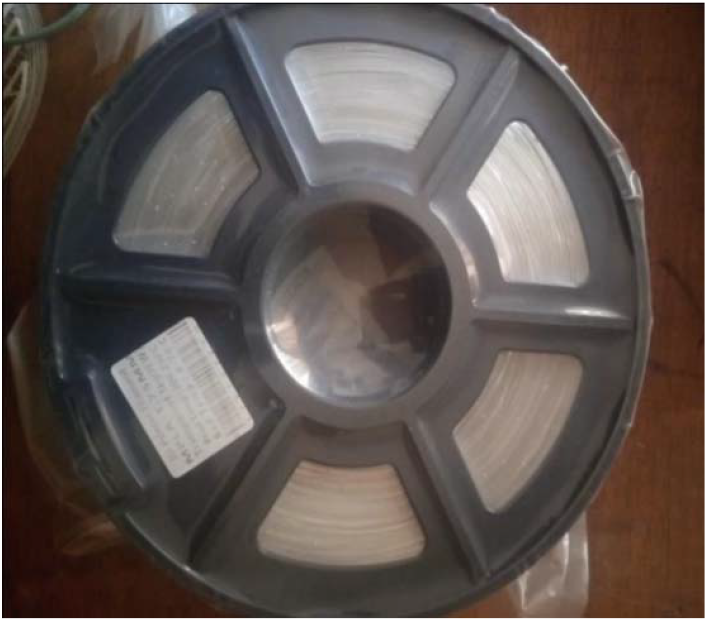
1.75mm PLA filamen.

**Fig. 3.**
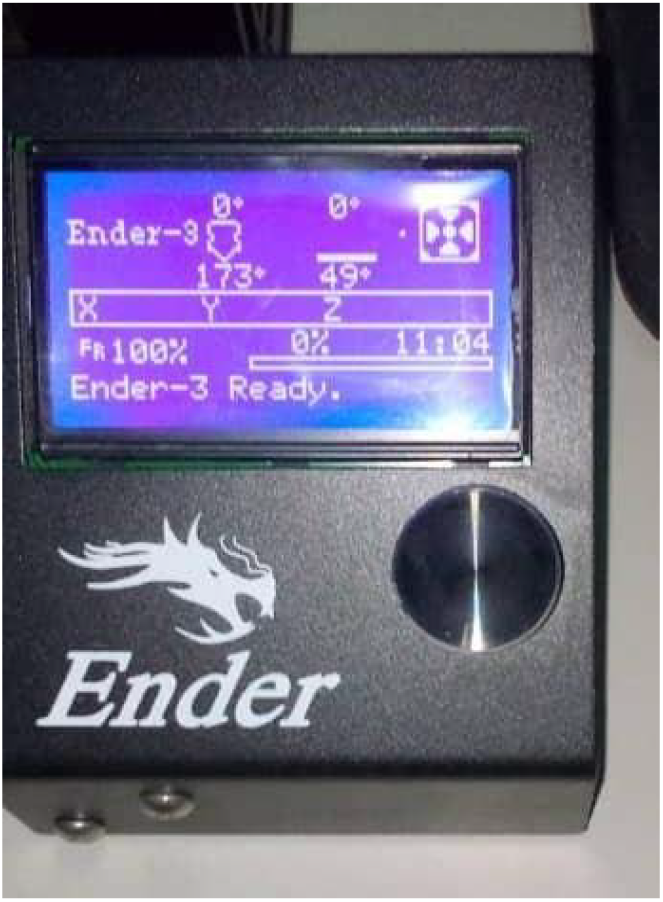
Ender-3 ready to print.

**Fig. 4.**
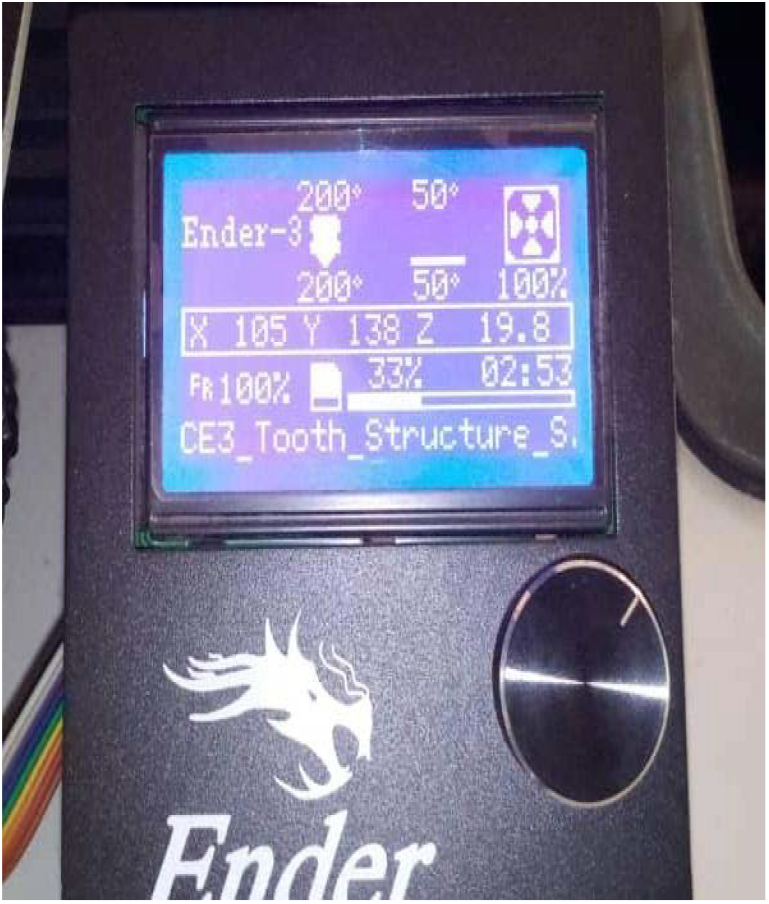
Ender-3 printing.

**Fig. 5.**
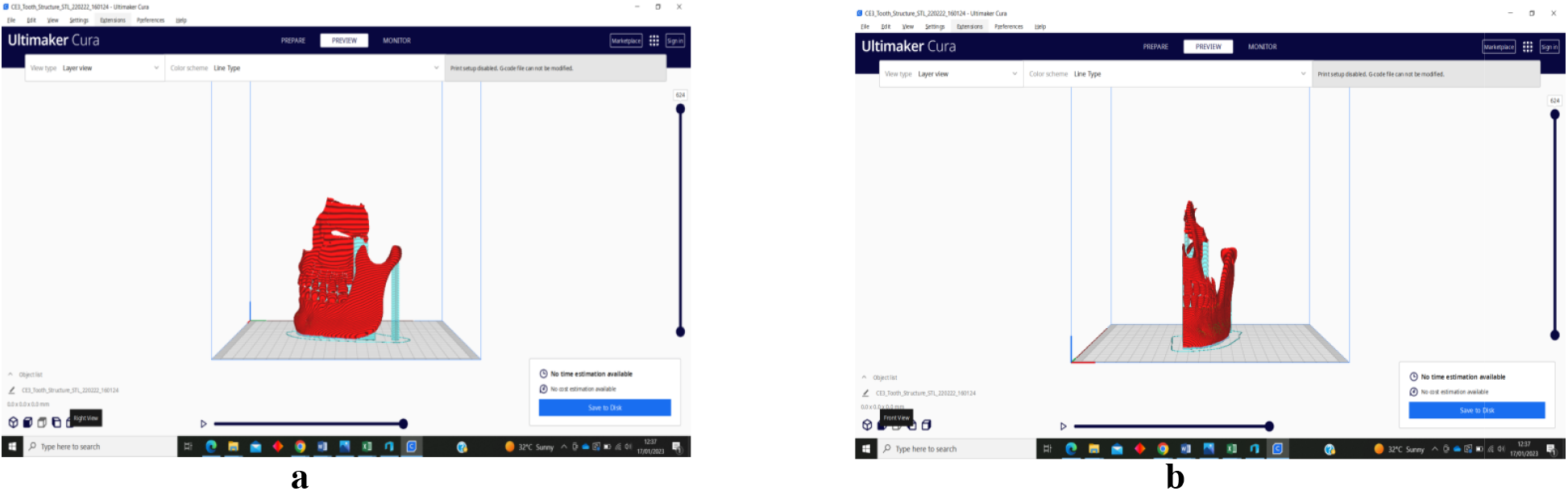
Ultimaker Cura rendering (a) left lateral view (b) anterior view.

**Fig. 6.**
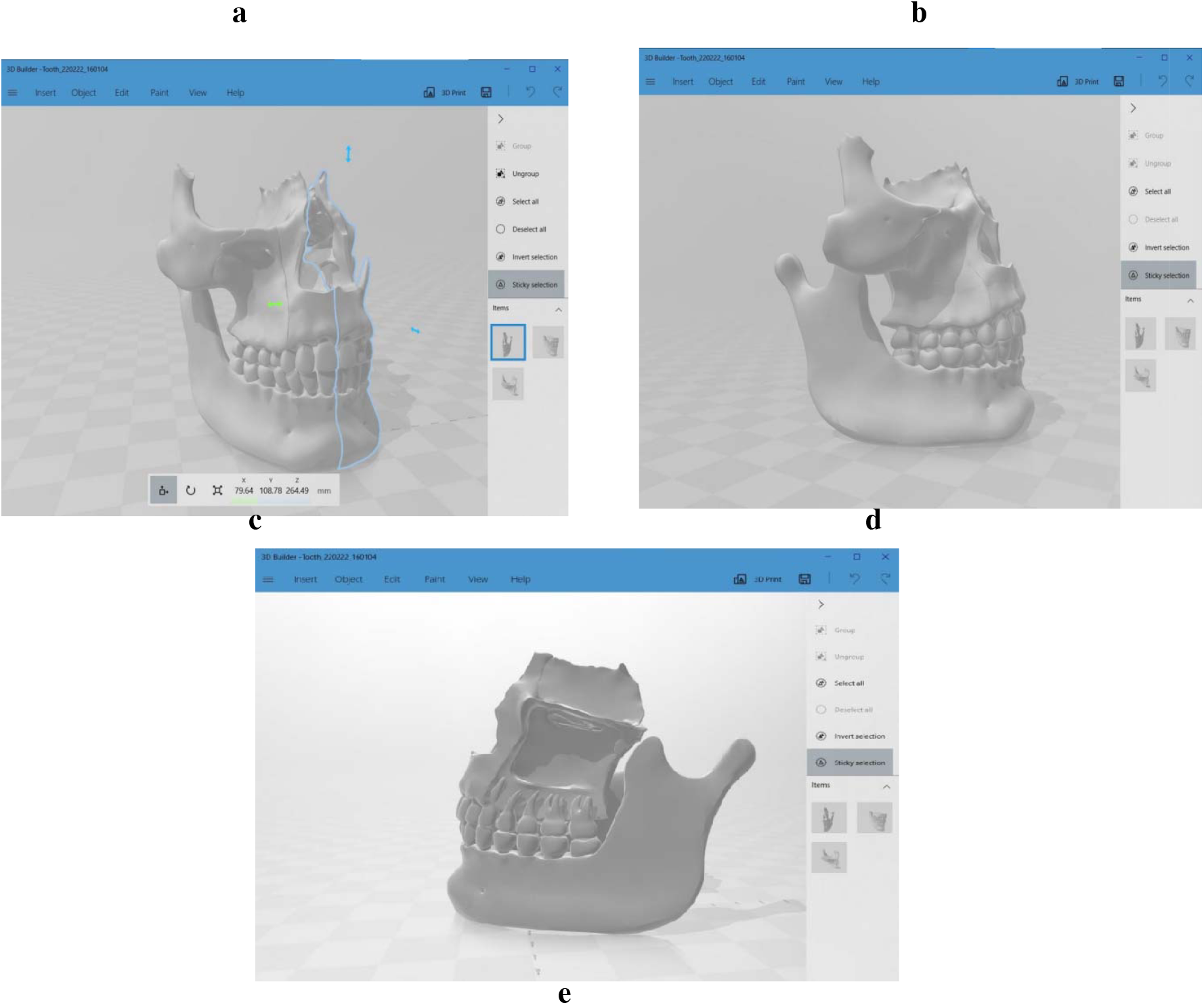
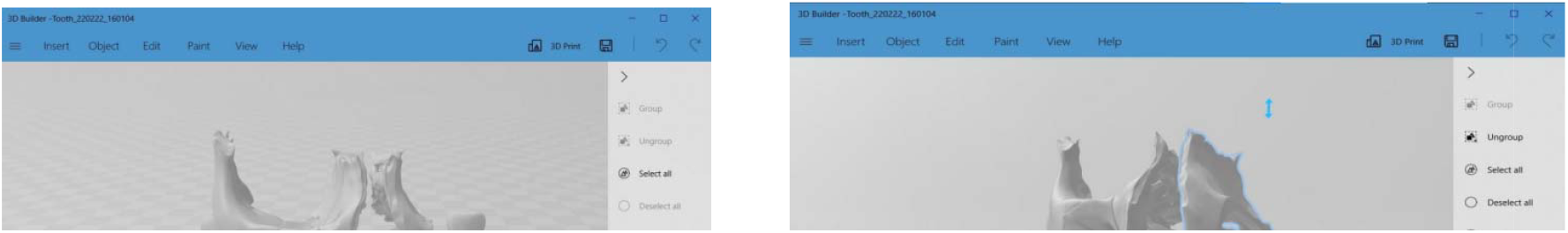
3D Builder rendering (a) anterior view (b) left rotation (c) right rotation (d) right la eral view (e) left lateral view.

Dimensional accuracy was verified to within **±0.3 mm** of the digital model using digital calipers.

**Fig. 7.**
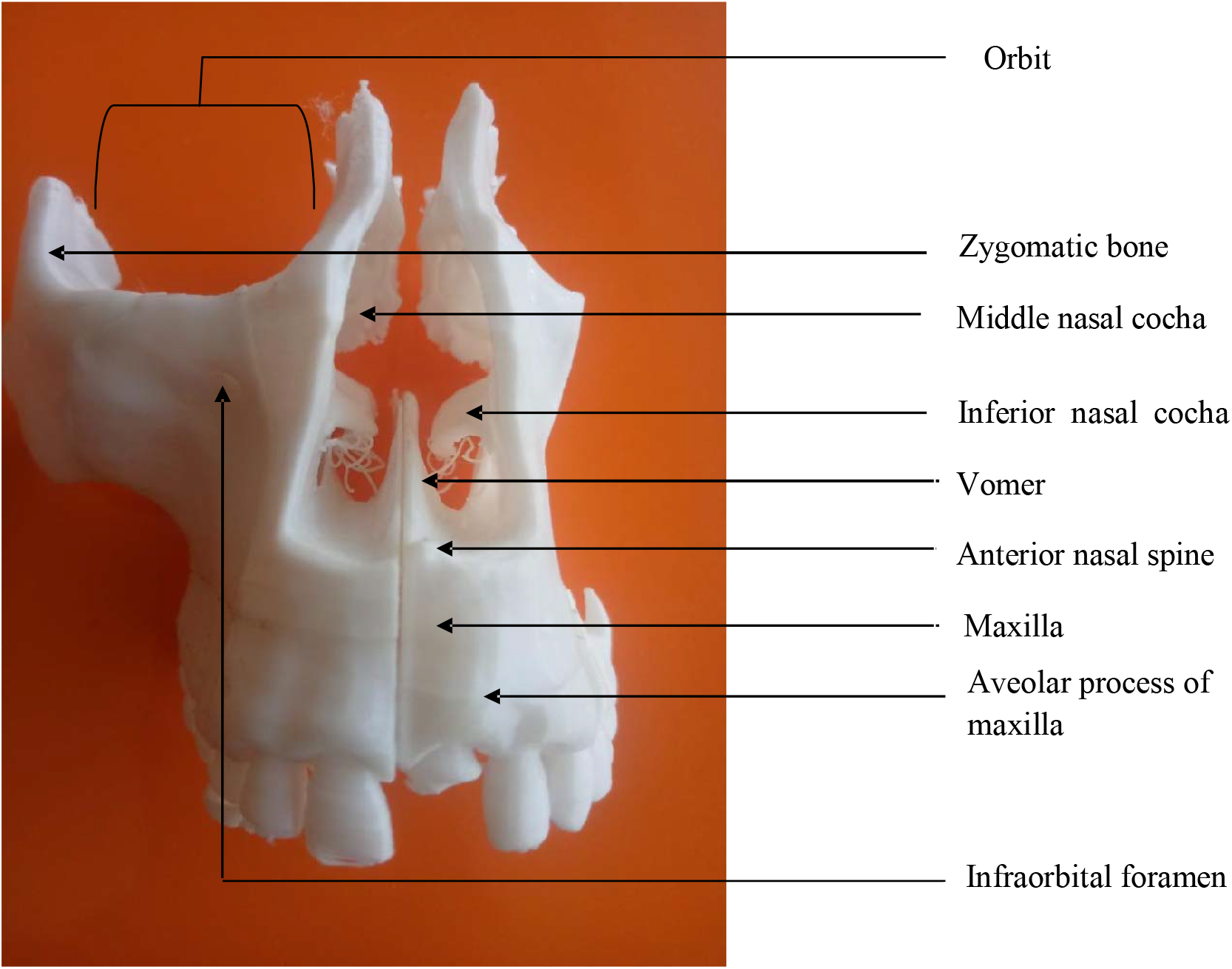
Aosterior side.

**Fig. 8.**
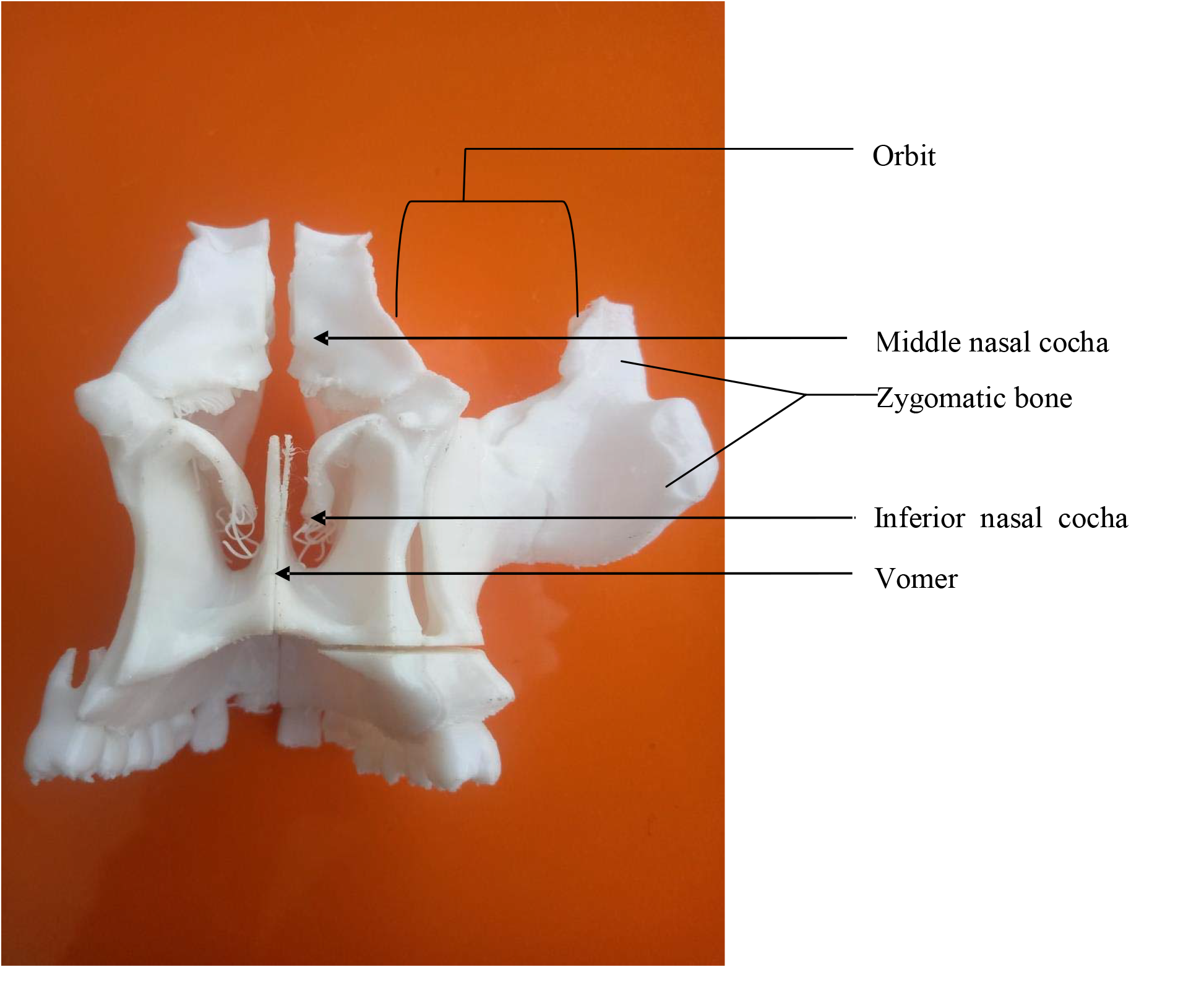
Posterior side.

**Fig. 9.**
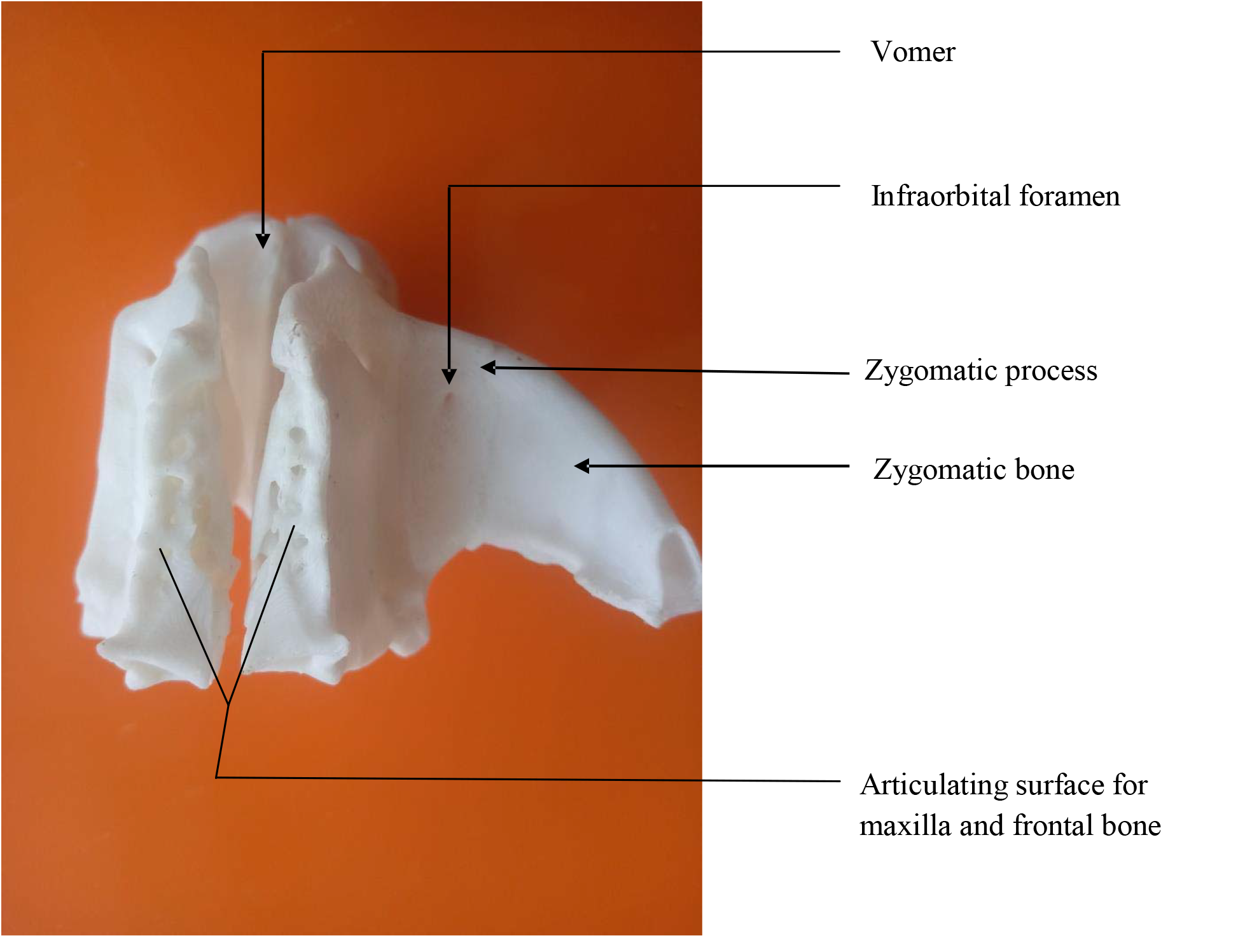
Superior side.

**Fig. 10.**
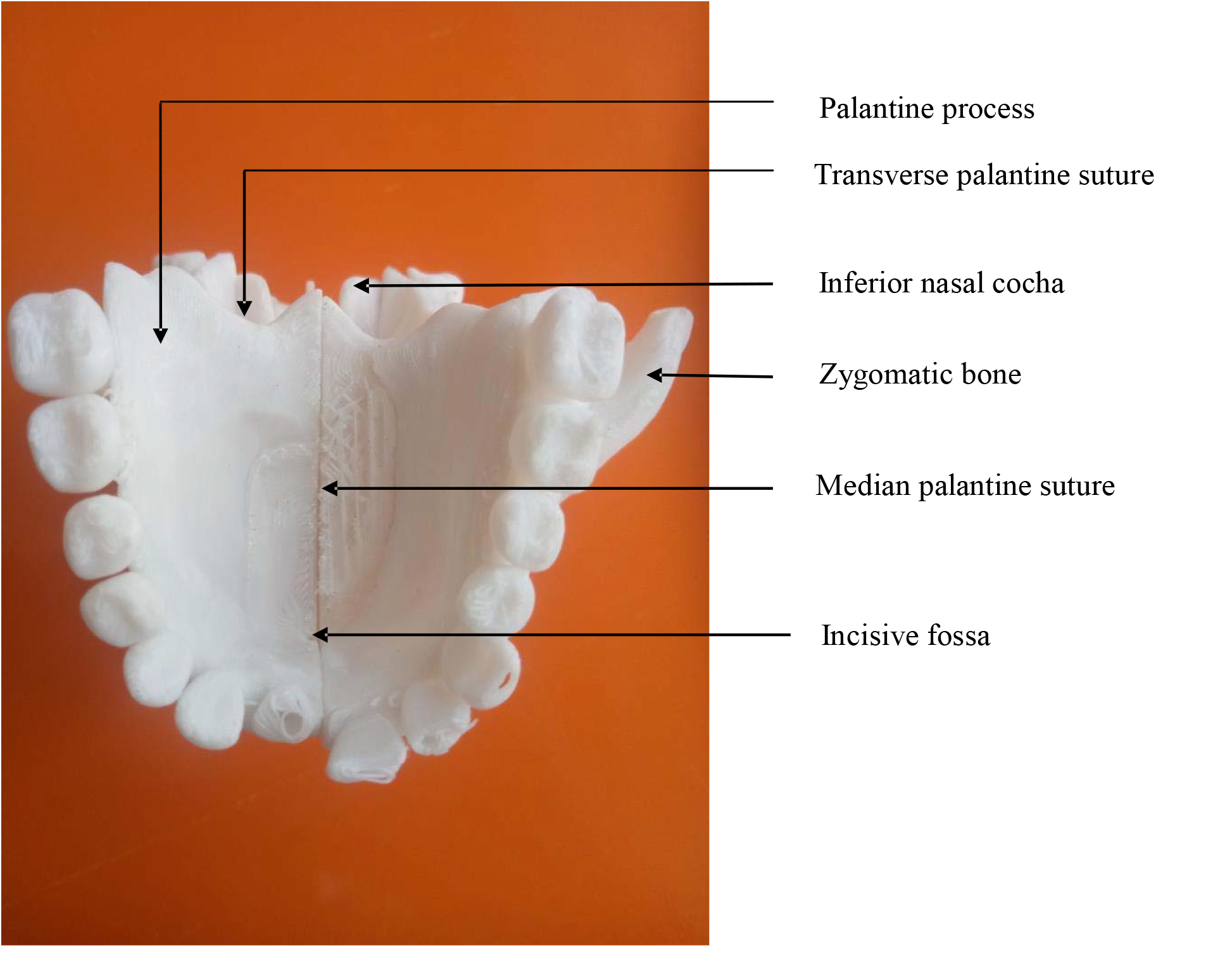
Inferior side.

**Fig. 11.**
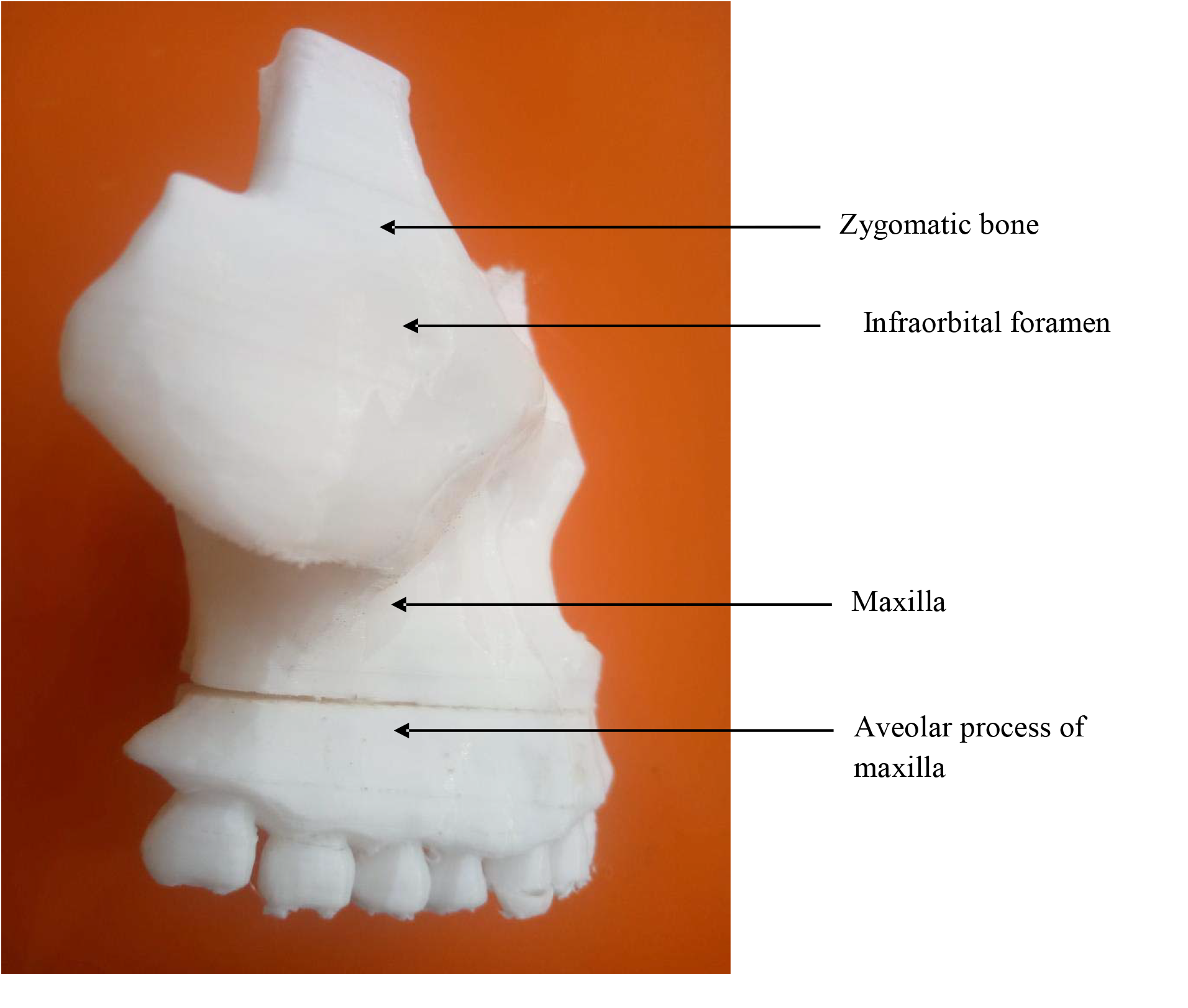
Right lateral side.

**Fig. 12.**
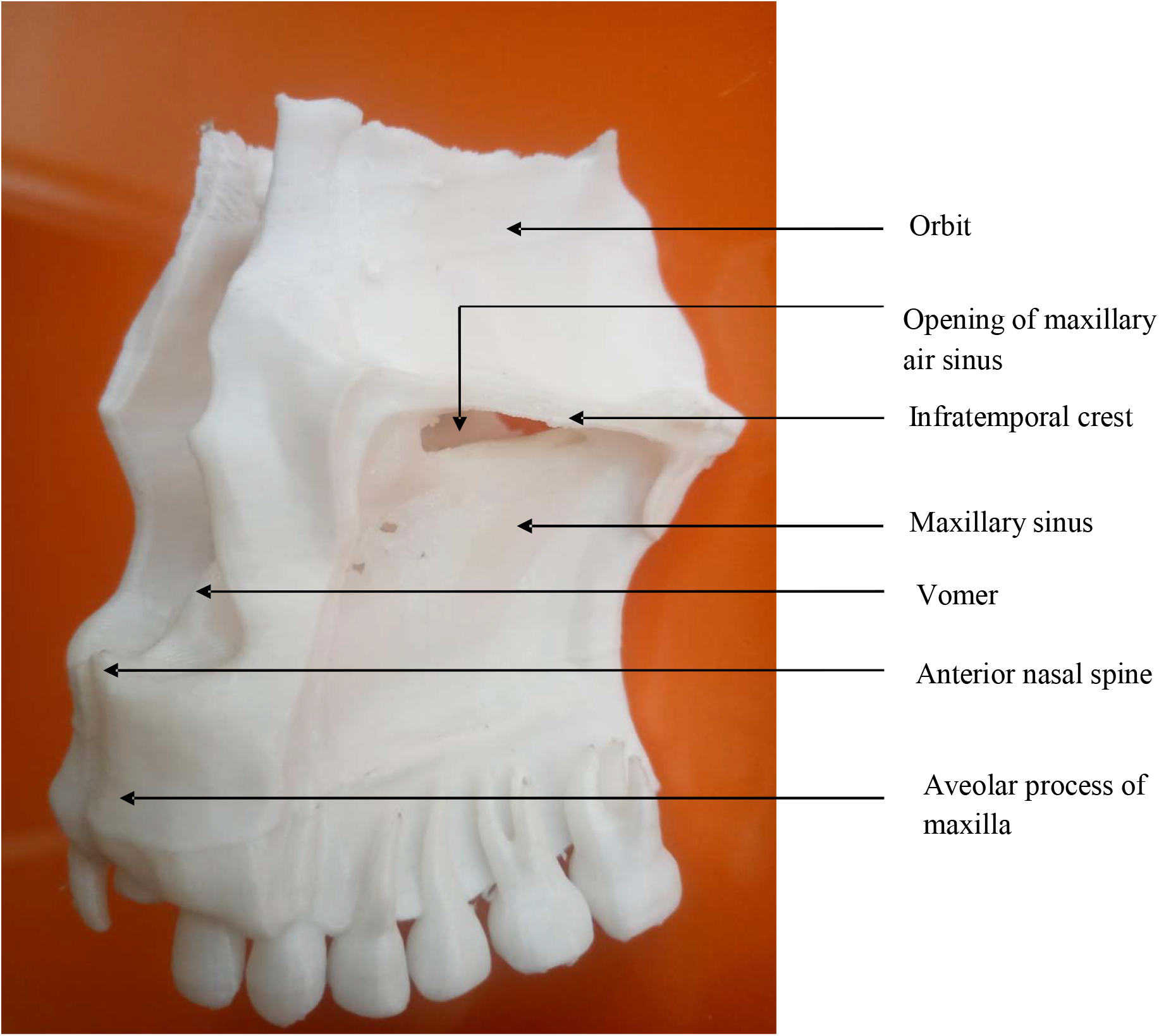
Left lateral side.

## DISCUSSION

The study successfully demonstrated the feasibility of producing anatomically accurate maxillofacial models using low-cost FDM 3D printing.

Key advantages include:

1. **Educational Value:** Models provide a tactile learning experience without the ethical concerns associated with cadaver use.
2. **Cost Efficiency:** Total material cost was **<$5 per model**, significantly lower than commercial anatomical models.
3. **Reproducibility:** Digital files enable unlimited identical reproductions, ensuring consistency in educational tools.

### Limitations

Surface resolution was limited by the **0.4 mm nozzle diameter**.
Anisotropic mechanical properties of PLA may affect model durability.
Color uniformity challenges arose due to the single-extruder system.

### Future Directions

– Explore **multi-material printing** for tissue differentiation.
– Incorporate **pathological variants** to enhance clinical relevance.
– Develop **regional anatomical model repositories** for resource-limited settings.

## CONCLUSION

This study highlights the potential of **low-cost 3D printing** to produce accurate, portable, and ethically compliant anatomical models. The approach is particularly valuable in **resource-limited settings**, such as sub-Saharan Africa, where access to cadavers is restricted. Future research should focus on refining printing techniques and expanding model applications to further enhance anatomical education.

## Data Availability

All data produced in the present study are available upon reasonable request to the authors.

## Notes

### Competing Interest Statement

The authors have declared no competing interest.

### Funding Statement

This study did not receive any funding.

